# Collaborative large language models (LLMs) are all you need for screening in systematic reviews

**DOI:** 10.64898/2026.02.07.26345640

**Authors:** Mihir Parmar, Syed Arsalan Ahmed Naqvi, Kainat Warraich, Amir Saeidi, Samarth Rawal, Kunwer Sufyan Faisal, Syeda Zainab Kazmi, Maurish Fatima, Huan He, Mohammad Safdar, Wei Liu, Tufia Haddad, Zhen Wang, Mohammad Hassan Murad, Chitta Baral, Irbaz Bin Riaz

## Abstract

**Background:** The ability of large language models (LLMs) to work collaboratively and screen studies in a systematic review (SR) is under-explored. Hence, we aimed to evaluate the effectiveness of LLMs in automating the process of screening in systematic reviews.

**Methods:** This is an observational study which included labeled data (title and abstracts) for five SRs. Originally, two reviewers screened the citations independently for eligibility. A third reviewer cross-checked each citation for quality assurance. GPT-4, Claude-3-Sonnet, and Gemini-Pro-1.0 were used using zero-shot chain-of-thought prompting. Collaborative approaches included (i): conflict resolution using benefit of the doubt, (ii) majority voting using an independent third LLM and (iii) conflict resolution using an informed third LLM. Performance was assessed using accuracy, precision for exclusion, and recall for inclusion. Work saved over samples (WSS) was computed to estimate the reduction in manual human effort.

**Results:** A total of 11300 articles were included in this study. The individual models, GPT-4, Claude-3-Sonnet, and Gemini-Pro-1.0 exhibited a high precision for exclusion, achieving 99.7%, 99.7%, and 99.2% and high recall for inclusion achieving 95.5%, 96.6% and 85.7%, respectively. However, the collaborative approach utilizing the two best-performing models (GPT-4 and Claude-3S) achieved an average precision of 99.9% and a recall of 98.5% (across all collaborative approaches). Furthermore, the proposed collaborative approach resulted in an average WSS of 63.5%, compared to the average WSS of 45.2% for individual models. Conversational LLM interactions showed a consistent pattern of results.

**Limitations:** This study was limited due to reliance on proprietary models, and evaluation on oncology datasets.

**Conclusion:** Evidence shows that collaborative LLMs enable efficient, high-performing screening in systematic reviews, supporting continuous evidence updates.

**Primary funding source:** NIH (U24CA265879-01-1) and Carolyn-Ann-Kennedy-Bacon Fund.

## INTRODUCTION

Systematic reviews form the foundation of evidence-based clinical practice and health policy, but their production involves substantial resource burdens. The screening phase represents a particularly labor-intensive bottleneck in the systematic review process. On average, screening titles and abstracts takes 33 days of analyst time, with reviewers often required to manually evaluate thousands of articles (1). The process is not merely time-consuming but vulnerable to human limitations. Reviewer fatigue can lead to errors and inconsistencies that potentially compromise review quality (2). Additionally, recent methodological studies have found that even dual independent screening—considered the gold standard—is imperfect, with single screeners missing a median of 5% of includable studies (3). The substantial time investment required for screening creates significant delays between research publication and evidence synthesis, limiting the timeliness and utility of systematic reviews for clinical and policy decision-making.

Given these challenges, there has been growing interest in facilitating the process of screening articles using machine learning (ML) models(4, 5). While traditional ML approaches have shown promise in assisting with large-scale screening tasks, they often rely on extensive training data, feature engineering, and lack the flexibility needed to generalize across various clinical questions. More recently, the advent of large language models (LLMs), has introduced a new frontier for automation. These models can process and understand vast amounts of unstructured text, offering the potential to revolutionize the screening process by leveraging their deep contextual understanding of the literature without task-specific fine-tuning (6).

Despite the potential of LLMs, current implementations typically assess their performance as standalone systems, akin to a single-reviewer approach (7). This contrasts with the conventional methodology of SRs, where two independent reviewers screen studies to minimize bias and improve accuracy (8). The single-reviewer model employed by LLMs lacks this critical collaborative aspect, which is fundamental in the real-world application of SRs. Thus, an important gap exists in current research: how well can LLMs perform when simulating the two-reviewer model commonly used in practice?

These limitations underscore the need for a more nuanced framework using LLMs, where they operate collaboratively to simulate the two-reviewer process. The purpose of this study is to evaluate the effectiveness of LLMs in automating the process of screening in systematic reviews. Specifically, we assessed the performance of collaborative LLM approaches, leveraging multiple LLMs as independent reviewers. We introduced and evaluated three distinct methods for conflict resolution between LLMs, reflecting real-world scenarios where two reviewers may disagree on the inclusion or exclusion of studies. Additionally, we explore the development of a conversational system in which LLMs simulate expert dialogue to resolve conflicts in the screening process.

## METHODS

This study is reported in accordance with the TRIPOD-LLM reporting guideline for studies using large language models

### Data sources

#### Dataset

The datasets from the living interactive evidence synthesis project were used for the analyses (9). Data from five ongoing systematic reviews were included focusing on: (i) systemic treatment options in metastatic hormone-sensitive prostate cancer (mHSPC) (10), (ii) systemic treatment options in metastatic castration-resistant prostate cancer (mCRPC) (11), (iii) systemic treatment options in renal cell carcinoma (RCC) (12), (iv) systemic treatment options in advanced/metastatic hepatocellular carcinoma (mHCC) (13), and (v) toxicity of PARP inhibitors in cancer patients.

#### Manual annotation

Citations were presented in a web-assisted graphical user interface where two independent reviewers screened citations based on prespecified inclusion/exclusion criteria for potential eligibility during the process of conducting the systematic review. The title and abstract of each citation are screened, and each citation is labeled based on the decision for potential inclusion or exclusion. Additionally, a third reviewer cross-checked each article confirming potential eligibility for this study. Details are provided in **Supplement Methods**.

#### Data processing

The user-generated labels, along with the corresponding meta-data, and free text for each stage of study selection were organized. Decision labels were transformed into a single binary variable with two levels: included (included by full-text review) and excluded (excluded by title/abstract or excluded by full-text review).

### Inference experiments

The process of screening was formulated as a generative task approach. Each title and abstract pair were evaluated based on predefined population, intervention, control, outcome, study design (PICOS) criteria and the article were included if these were met.

#### Prompt development

A sample of 50 articles were used for iterative prompt development. A zero-shot chain-of-thought prompt (14) without specific in-context or additional examples was developed. The prompt was structured as (1), where *p* is the natural language prompt consisting of the task, inclusion criteria and output format, *t* is the title, and *a* is the abstract for a given article. The prompts for different experimental setups are provided in **Supplement Tables 1-3.**

#### Models

Three commercial (closed-source) LLMs (GPT-4 Turbo [4T], Claude-3 Sonnet [3S], Gemini-Pro-1.0 [P]) were used utilizing the versions released in May 2024. The temperature hyperparameter was set to be 0 ensuring deterministic responses with a maximum output of 1,000 tokens.

### Experimental setups

#### Independent individual large language models

Using the same prompts as described above, an explanation and a final decision for each given article were generated at the level of each of the three LLMs independently as shown in **Supplement Figure 1**.

#### Collaborative large language models with conflict resolution

A collaborative decision-making process involving two LLMs, GPT-4T and Claude-3S as reviewers was employed using the same zero-shot chain-of-thought prompt. Each article is screened independently by the two LLMs (GPT-4T and Claude-3S). The explanation and decision are generated at the level of each article. Subsequently, if there is concordance between the decisions of the two LLMs, the decision is finalized. In cases of discrepancies or discordance between the two reviewers (LLMs), conflicts were resolved using the following three approaches (**Figure 1**):

i. *Conflict resolution using benefit of the doubt (BoD approach):* The article(s) is included given that there was a discrepancy in the decision of the two LLMs. This gives the BoD to the article and simulates the real-world practice of including articles for further review by full-text in cases of uncertainty.
ii. *Majority voting using an independent third LLM (MV approach):* The conflicts are resolved by an independent input from a third LLM (Gemini-P). The approach and the prompt used for calling Gemini-Pro-1.0 are the same as those used for GPT-4 and Claude-3-Sonnet. The explanation and decision are generated at the level of each conflict article. Finally, the decisions for a given article are compared across the three LLMs (GPT-4T(15), Claude-3S(16), Gemini-P(17)) and the most common decision is selected usingMV.
iii. *Conflict resolution using an informed input from a third LLM (Informed Input approach):* The conflicts are resolved using an informed input from a third LLM (GeminiP) by providing the conflicted article, and conflicting decisions along with the explanations from the two LLMs (GPT-4T and Claude3S). After reviewing the initial outputs from the two LLMs, Gemini-P generates a final decision with an explanation. This ensures an approach that is informed by prior outputs to generate a finalized decision leveraging the ability of LLMs to critique.

**Figure 1:**
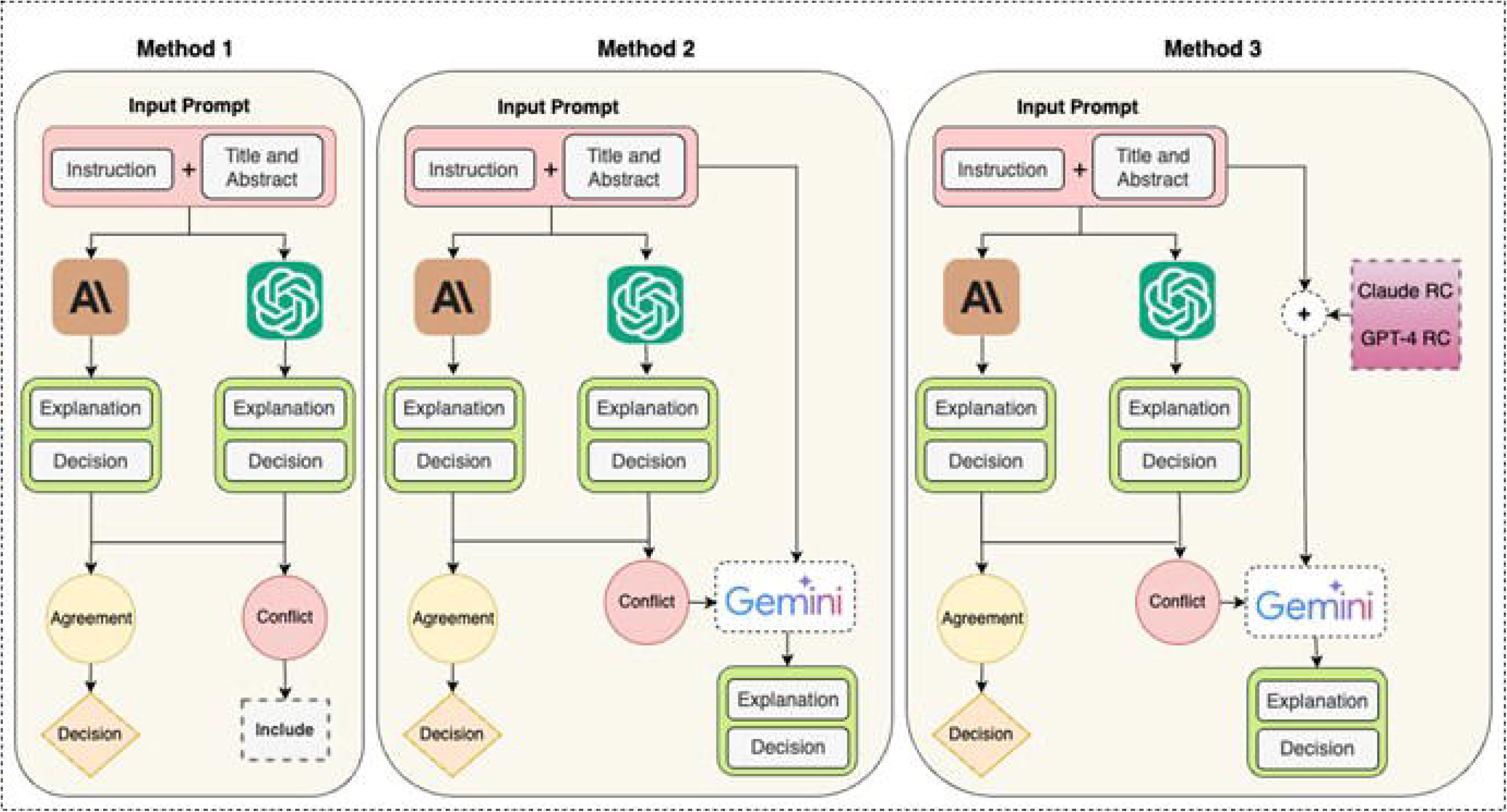
A schematic representation of collaborative LLMs approaches for conflict resolution. We evaluated three different approaches: (i) Benefit of Doubt, (ii) Majority Voting, and (iii) Informed input. In the “Benefit of Doubt” approach (Method 1 in Figure 1), any conflict between two models is resolved by automatically labeling the article “Include.” In the “Majority Voting” approach (Method 2 in Figure 1), we query a third model and assign the label supported by at least two of the three models by majority voting. In the “Informed Input” approach (Method 3 in Figure 1), we present both conflicting decisions and their reasoning chains from the first two models to a third model, which then makes a final, informed labeling decision.

#### Multi-large language model conversational interaction

Additionally, a post-hoc case study was conducted to simulate the conversation of two experts using the best-performing models, GPT-4T and Claude-3S in an attempt to reach a final decision for a given study. We used conflicted samples (between the two LLMs) from the mCRPC dataset where the overall recall for inclusion was relatively lower to assess the additive utility of this approach. Details of cross-LLM conversational set up are provided in **Supplement Figure 2,** and **Supplement Tables 3-4.**

### Performance evaluation

The performance of LLMs in each experimental setup was assessed using precision, recall, and overall accuracy. We reported precision for exclusion and recall for inclusion, ensuring that the models were evaluated based on their ability to precisely exclude irrelevant articles while not missing articles meant to be included.

### Statistical analysis

Descriptive statistical analysis were conducted; categorical variables were summarized as frequencies with relative percentages and continuous variables were summarized as mean with standard deviations. Work saved (WS) (18) was computed for each systematic review across different experimental setups to quantify the reduction in manual-human-effort as:

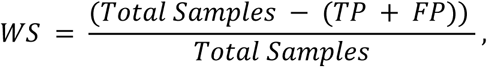

where “Total Samples” refers to the total number of articles that need to be reviewed manually for each dataset, “TP” true positives, and “FP” represents false positives.

Furthermore, the work saved over sample (WSS) was computed by normalizing WS by the number of missing included articles (i.e., loss) as follows:

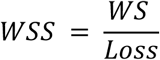

A higher normalized WS value would indicate a model that not only has a high WS but also a low loss (indicating fewer missed relevant articles). Conversely, a lower normalized WS value would suggest that the model, despite having a reasonable WS, misses a significant number of relevant articles. Hence, normalizing the value of WS by loss (i.e., WSS) provides a measure that reflects the trade-off between the reduction in human effort (i.e., WS) and the number of relevant articles missing. This normalized value can be interpreted as an efficiency score, indicating the extent to which human effort has been reduced while minimizing the number of missed relevant articles.

### Error analysis

A systematic error analysis was performed by a trained systematic reviewer to assess the reasoning chain of each model’s decision for each article. Errors were first stratified by the population, intervention, control/comparator, and outcomes (PICOS) elements and then were further categorized using a pre-specified error categorization schema to differentiate errors that are likely due to comprehension from those that are likely due to reasoning as shown in **Supplement Figure 3**.

### Data availability

Source code and fully annotated datasets with gold-standard decision labels and machine-generated decisions labels with explanation are publicly available at https://github.com/Mihir3009/SR_LLMs.

## RESULTS

A total of 11300 articles across five prespecified systematic reviews were included in this study. Of the total, 10530 (93.2%) were labeled as excluded, and only 770 articles (6.8%) were labeled as included. The greatest number of articles were derived from the PARP toxicity systematic review (N = 4775; 42.2%), followed by mHCC (N = 1998; 17.7%), mCRPC (N = 1637; 14.5%), mHSPC (N = 1552; 13.7%), and RCC systematic review (N = 1338; 11.8%) as shown in **Table 1**.

**Table 1.** Characteristics of the included systematic review datasets.

|  | **mHSPC** | **mCRPC** | **RCC** | **mHCC** | **PARPi**<br>**toxicity** | **Total** |
| --- | --- | --- | --- | --- | --- | --- |
| **Total articles** | 1552 | 1637 | 1338 | 1998 | 4775 | 11300 |
| **Decision labels - N (%)** |  |  |  |  |  |  |
| Excluded | 1524<br>(98.19) | 1526<br>(93.22) | 1253<br>(93.65) | 1967<br>(98.45) | 4260<br>(89.21) | 10530<br>(93.19) |
| Included | 28<br>(1.8) | 111<br>(6.78) | 85<br>(6.35) | 31<br>(1.55) | 515<br>(10.79) | 770<br>(6.81) |

Abbreviations: N – number; mHSPC: metastatic hormone-sensitive prostate cancer; mCRPC: metastatic castration resistant prostate cancer; RCC: renal cell carcinoma; mHCC: metastatic hepatocellular carcinoma; PARPi: poly (ADP-ribose) polymerase inhibitors

### Performance

The overall performance for the three independent and individual LLMs (GPT-4T, Claude-3S, Gemini-P-1.0) and collaborative LLMs with conflict resolution across the five systematic reviews is outlined in **Table 2 and Figures 2-3**.

**Figure 2.**
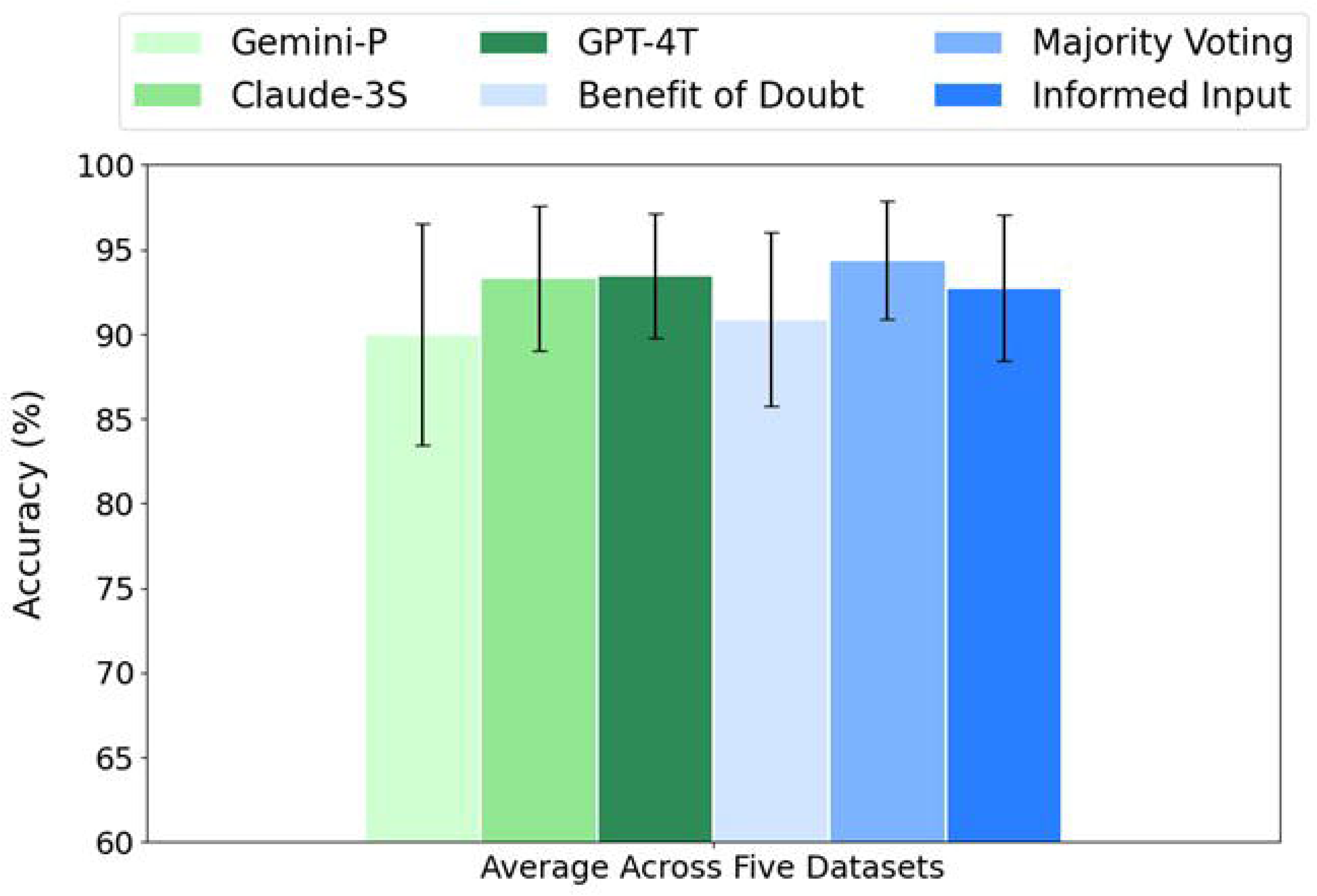
Average overall accuracy for independent LLMs as well as our proposed approaches.

**Figure 3.**
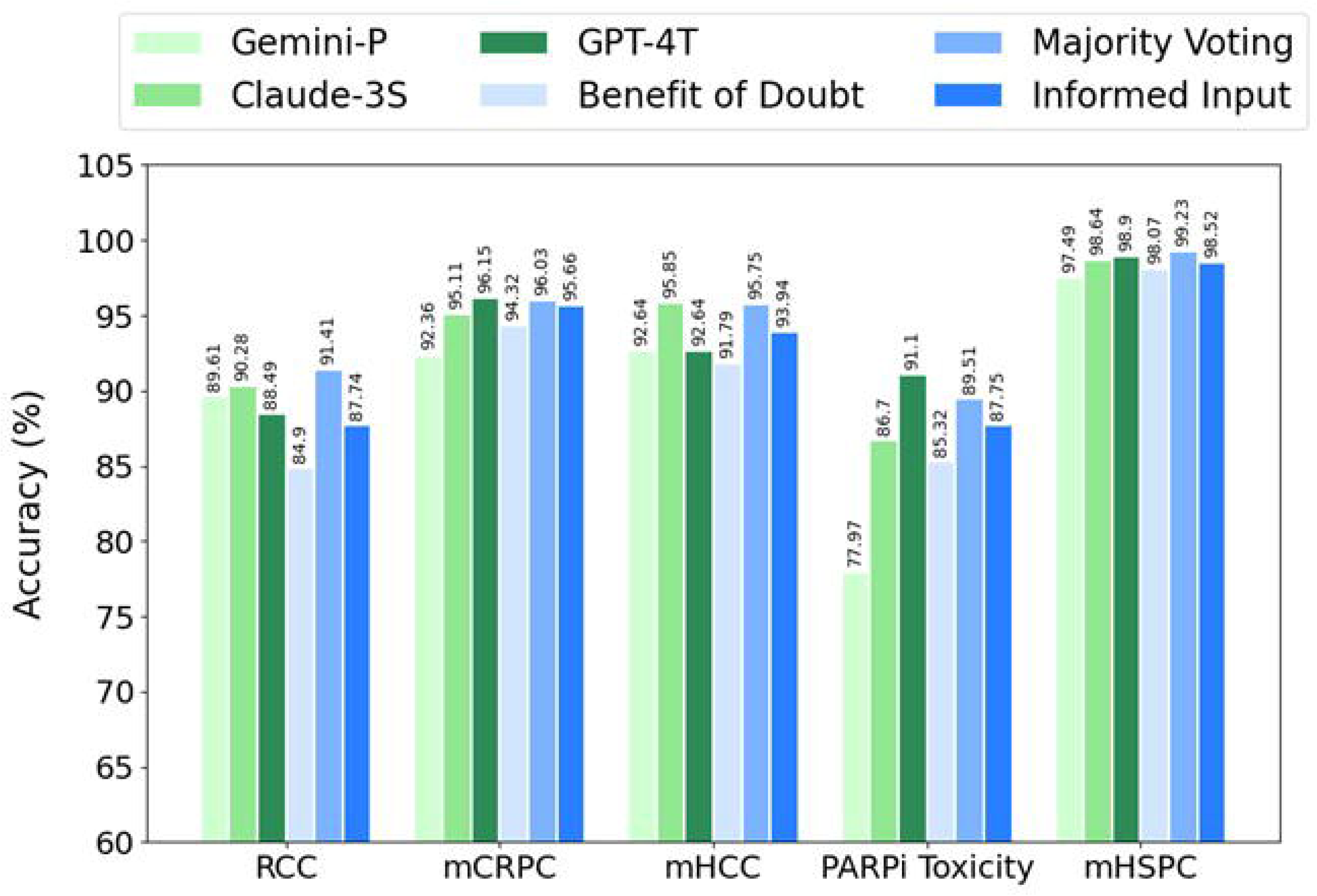
Evaluation of large language models in terms of overall accuracy across the five systematic review datasets for independent LLMs as well as our proposed approaches.

**Table 2.**
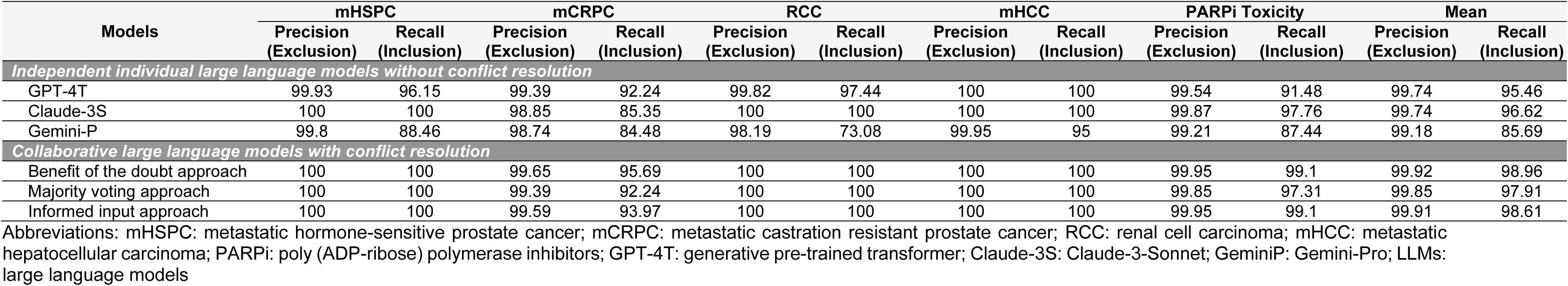
Performance of independent individual LLMs and collaborative LLMs with conflict resolution.

#### Independent individual LLMs without conflict resolution

Overall, GPT-4T exhibited a mean precision of 99.74 for excluding articles and a mean recall of 95.5% for including articles. Across systematic review projects, precision for excluding articles ranged from 99.4% in the mCRPC systematic review project to 100% in the mHCC systematic review. Recall for including articles ranged from 91.5% in PARP toxicity to 100% in mHCC.

Claude-3S exhibited a mean precision of 99.7% for exclusion and a mean recall of 96.62% for inclusion. Across systematic review projects, precision for excluding articles ranged from 98.8% in the mCRPC project to 100% in mHSPC, RCC, and mHCC. Recall for including articles ranged from 85.3% in mCRPC to 100% in mHSPC, RCC, and mHCC.

Gemini-P exhibited a mean precision of 99.2% for exclusion and a numerically lower mean recall of 85.7% for inclusion as compared to GPT-4T and Claude-3S. Across systematic review projects, precision for excluding articles ranged from 98.2% in the RCC project to 99.9% in mHCC. Recall for including articles ranged from 73.1% in RCC to 95% in mHCC.

#### Collaborative LLMs with conflict resolution

Overall, the *BoD* approach exhibited a mean precision of 99.9% for excluding articles and a mean recall of 99% for including articles. Across systematic review projects, precision for excluding articles ranged from 99.7% in mCRPC to 100% in multiple reviews, including mHSPC, RCC, and mHCC. Recall for including articles ranged from 95.7% in mCRPC to 100% in mHSPC, RCC, and mHCC.

The *MV* approach exhibited a mean precision of 99.8% for excluding articles and a mean recall of 97.9% for including articles. Precision for excluding articles ranged from 99.4% in mCRPC to 100% in mHSPC, RCC, and mHCC. Recall for including articles ranged from 92.2% in mCRPC to 100% in mHSPC, RCC, and mHCC.

The *Informed Input* approach exhibited a mean precision of 99.9% for excluding articles and a mean recall of 98.6% for including articles. Precision for excluding articles ranged from 99.6% in mCRPC to 100% in mHSPC, RCC, and mHCC. Recall for including articles ranged from 94% in mCRPC to 100% in mHSPC, RCC, and mHCC.

### Manual work saved

The overall WS, and WSSlost for the three independent and individual LLMs (GPT-4T, Claude-3S, Gemini-P-1.0) and collaborative LLMs with conflict resolution across the five systematic reviews are outlined in **Table 3**. An average WSS performance across three individual LLMs is 45.2%, and across three collaborative approach is 63.5%.

**Table 3.** Work saved and work saved over samples lost across approaches.

| Models | mHSPC |  |  | mCRPC |  |  | RCC |  |  | mHCC |  |  | PARPi Toxicity |  |  | Mean |  |  |
| --- | --- | --- | --- | --- | --- | --- | --- | --- | --- | --- | --- | --- | --- | --- | --- | --- | --- | --- |
|  | Loss | WS | WSS | Loss | WS | WSS | Loss | WS | WSS | Loss | WS | WSS | Loss | WS | WSS | Loss | WS | WSS |
| *Independent individual large language models without conflict resolution* |  |  |  |  |  |  |  |  |  |  |  |  |  |  |  |  |  |  |
| GPT-4T | 1/26 | 97.4 | 97.4 | 9/116 | 90.2 | 10 | 2/78 | 83 | 41.5 | 0/20 | 91.6 | 91.6 | 19/223 | 87.2 | 4.59 | 31/463 | 89.9 | 49 |
| Claude-3S | 0/26 | 97 | 97 | 17/116 | 90.1 | 5.3 | 0/78 | 84.5 | 84.5 | 0/20 | 94.9 | 94.9 | 5/223 | 82.2 | 16.5 | 22/463 | 89.7 | 59.6 |
| Gemini-P | 3/26 | 96.3 | 32.1 | 18/116 | 87.5 | 4.86 | 21/78 | 87 | 4.14 | 45311 | 91.7 | 91.7 | 28/223 | 74.5 | 2.66 | 71/463 | 87.4 | 27.1 |
| *Collaborative large language models with conflict resolution* |  |  |  |  |  |  |  |  |  |  |  |  |  |  |  |  |  |  |
| Benefit of the doubt approach | 0/26 | 96.4 | 96.4 | 5/116 | 87.4 | 17.5 | 0/78 | 79.1 | 79.1 | 0/20 | 90.8 | 90.8 | 2/223 | 80.6 | 40.3 | 7/463 | 86.8 | 64.8 |
| Majority voting approach | 0/26 | 97.6 | 97.6 | 9/116 | 89.3 | 9.92 | 0/78 | 85.6 | 85.6 | 0/20 | 94.8 | 94.8 | 6/223 | 84.7 | 14.1 | 15/463 | 90.4 | 60.4 |
| Informed input approach | 0/26 | 96.9 | 96.9 | 7/116 | 88.8 | 12.7 | 0/78 | 82.1 | 82.1 | 0/20 | 93.1 | 93.1 | 2/223 | 83.2 | 41.6 | 9/463 | 88.8 | 65.3 |

Abbreviations: mHSPC: metastatic hormone-sensitive prostate cancer; mCRPC: metastatic castration resistant prostate cancer; RCC: renal cell carcinoma; mHCC: metastatic hepatocellular carcinoma; PARPi: poly (ADP-ribose) polymerase inhibitors; GPT-4T: generative pre-trained transformer; Claude-3S: Claude-3-Sonnet; GeminiP: Gemini-Pro; LLMs: large language models; Loss: false exclusion of articles that were meant to be included; WS: work saved; WSS: work saved over samples lost
\*A higher work saved (WS) indicates that the model/approach is more effective in reducing manual screening efforts by screening a large portion of articles. For example, WS value of 89.87 would indicate that for every 100 articles screened manually, the model effectively screened out 89.87 the articles, resulting in only about 10.13% needing manual review. The work saved over samples lost (WSS) normalizes WS by considering how many relevant articles (articles that were meant to be included) were missed (loss). For example, WSS value of 49.02 would indicate that for every 100 articles screened manually, the model effectively screened out 49.02 articles at the expense of one article loss

#### Independent individual LLMs without conflict resolution

Overall, GPT-4T exhibited a mean loss of 31 articles with a WS of 89.9% and a WSS of 49%. Across systematic review projects, the WS ranged from 83% in the RCC project to 91.6% in mHCC.

Claude-3S exhibited a mean loss of 22 articles, achieving a WS of 89.7% and a WSS of 59.6%. The WS varied from 84.4% in the RCC project to 94.8% in mHCC.

Gemini-P exhibited a mean loss of 71 articles, with a WS of 87.4% and a WSS of 27.1%. The WS ranged from 87% in the RCC project to 91.7% in mHCC.

#### Collaborative LLMs with conflict resolution

Overall, the BoD approach exhibited a mean loss of 7 articles with a WS of 86.8% and a WSS of 64.8%. The WS across systematic review projects ranged from 79.1% in mHCC to 90.8% in mHSPC.

The MV approach exhibited a mean loss of 15 articles, with a WS of 90.4% and a WSS of 60.4%. The WS ranged from 89.2% in mCRPC to 94.8% in mHSPC.

The *Informed Input* approach exhibited a mean loss of 9 articles, achieving a WS of 88.8% and a WSS of 65.3%. The WS varied from 82.1% in mHCC to 93.1% in mHSPC.

### Conversational LLM-LLM interaction

Using the conflicted articles in the mCRPC dataset, the post-hoc analysis showed that the precision for inclusion was increased to 55.2% with the conversational cross LLMs interaction approach as compared to 45.1% (*BoD* approach), 54.2% (*MV*approach), and 52.6% (*Informed Input* approach). However, the recall for inclusion was decreased to 87.7% with the conversational cross LLMs interaction approach as compared to 94.3% (*BoD*approach), 89.3% (*MV*approach), and 91% (*Informed Input* approach). An example of a conversation between GPT-4T and Claude-3S in which they had a conflict is presented in **Supplement Tables 3-4**.

### Error analysis

A total of 517 studies (22.7%) were incorrectly categorized by GPT-4T, with 41 studies (7.9%) falsely excluded and 476 studies (92.1%) were falsely included. A total of 490 studies (21.5%) were incorrectly categorized by Claude-3S with 21 studies (4.3%) falsely excluded and 469 studies (95.7%) were falsely included (95.7%). A total of 1270 studies (55.8%) were incorrectly categorized by Gemini-P with 102 studies (8.0%) falsely excluded, and 1168 studies (91.7%) were falsely included. A detailed breakdown by each systematic review data set is provided in **Supplement Table 5.**

The PICOS component analysis, the largest proportion of errors were related to study design with GPT-4T (N = 364; 70.4%), and to intervention with both Gemini-P (N = 717; 57.5%) and Claude-3S (N = 231; 48.9%).

In terms of comprehension and reasoning errors, all three models displayed a high proportion of comprehension errors (GPT-4T: N = 496, 95.9%; Claude-3S: N = 449, 91.6%; Gemini-P: N = 1216; 95.7%) Detailed results are summarized in **Supplement Tables 6-9.**

## DISCUSSION

The current study evaluated the performance of three LLMs; GPT-4T, Claude-3S, and Gemini-P) for screening articles across five systematic reviews. Our findings demonstrate that while individual models achieved high precision and effective recall, the proposed collaborative approaches - simulating two reviewer settings in real world – resulted in a substantial increase in overall performance, with mean precision reaching as high as 99.9% and recall as high as 99%. All three collaborative LLMs approaches demonstrated numerically better performance with substantial precision for exclusion and recall for inclusion on three systematic reviews with considerable increase in recall for inclusion on the remaining two systematic reviews datasets. In terms of individual LLMs without conflict resolution, Gemini-P model, although precise for excluding articles (mean precision ∼98%), showed comparatively lower recall for inclusion (mean recall 84%), suggesting a higher likelihood of missing relevant articles. While GPT-4T and Claude-3S models also performed well individually, they still fell short of the proposed BoD and *MV* approaches. This collaborative framework also reduced potential manual screening effort by approximately 60%. Additionally, our case study on LLM-LLM conversational interaction suggests this could potentially be used to further improve the performance, potentially attenuate manual workload, and make more informed and transparent screening decisions using LLMs.

In contrast, previous efforts to automate the title and abstract screening process have typically relied on labor- or computationally intensive methods, such as supervised labeling, pretraining, or vectorization (5). For example, Rayyan (19) and Abstrackr (20) are two free web tools that offer a semi-automated approach to article filtering using natural language processing (NLP) algorithms to learn from a reviewer’s decisions with subsequent replication of the process. Beyond these standard methods, past attempts have also been made to utilize pre-trained generative models. For instance, Syriani et al. (21) used ChatGPT for document screening and found that its effectiveness is limited when the document set is imbalanced — a common issue in systematic reviews. Bio-SIEVE (22), a model fine-tuned from the Guanaco checkpoint (built on Llama architecture), demonstrated higher classification accuracy than ChatGPT for article screening. Wang et al. (23) evaluated the effectiveness of eight LLMs by exploring a calibration technique that applies a predefined recall threshold to determine eligibility of a given article. However, these efforts have all relied on the capabilities of a single LLM. Conversely, the approach in this study is the first to simulate the real-world two-reviewer process by employing two independent LLMs. This innovative method not only mirrors the collaborative nature of human reviewers but also outperforms strategies that use a single LLM, setting a new benchmark in evaluating the performance of these models for screening articles.

Moreover, these findings have important implications for clinical practice guidelines. Clinical practice guidelines can lose relevance within two to three years of publication(24), necessitating frequent updates to ensure optimal patient care. This is especially pertinent in rapidly evolving fields such as oncology, where clinical guidelines can become outdated quickly due to the fast pace of evolving evidence and therapeutic innovation(25, 26). The formulation of a clinical guideline is a resource-intensive process, typically requiring months to years of effort from multidisciplinary teams(24, 27). A significant portion of this time is dedicated to conducting systematic reviews, with the screening phase alone often taking weeks to months. The results presented in this study suggest a paradigm shift in how systematic reviews could be conducted, particularly for living clinical practice guidelines. By demonstrating the high precision for exclusion and recall for inclusion with collaborative LLMs in screening titles and abstracts, the current findings open up possibilities for significantly accelerating the systematic review process. The collaborative LLM approach would potentially allow for more frequent and comprehensive updates to living guidelines. This coupled with the WSScould translate into faster turnaround times for guideline updates and more efficient resource allocation in the guideline development process.

While our approach leveraging collaborative LLMs shows significant promise in automating SRs, there are several limitations of this study. The dependency on proprietary LLMs, such as GPT-4T, Claude-3S, and Gemini-P, might reduce the reproducibility, as these models undergo frequent updates which might affect performance consistency. However, open-source models can be leveraged using the same proposed approach and would allow for greater control and customization. Although we used a random sample of articles to develop our prompts, it is possible that the models may have encountered these articles during their pre-training phase, potentially leading to an overestimation of their performance in a zero-shot setting. Moreover, it could be argued the use of zero-shot prompts can also result in variability due to the absence of specific in-context examples or fine-tuning tailored to each clinical question. Hence, the use of few-shot prompting or LLMs fine-tuned specifically for screening articles might offer even better performance. However, it should be noted that while a few in-context examples can be provided in large-scale systematic reviews, it may not be feasible to do so in smaller reviews where identifying “inclusion examples” would require substantial effort; at that point, it might be more practical to conduct the systematic review manually rather than search articles to be included as examples. Likewise, the relative proportion of included articles in the dataset was less compared to excluded articles; hence it is important to interpret the evaluation metrics (precision, recall) in context of the underlying prevalence of the eligible articles. Our method, though effective, involves computationally intensive processes, particularly during LLM-LLM interactions for conflict resolution, which can limit scalability in resource-constrained environments. Optimizing efficiency through techniques like model distillation or more computationally efficient architectures could make the approach more scalable and accessible. Lastly, despite achieving high precision and recall, these models still exclude a limited set of articles that met the inclusion criteria. Error analysis revealed that most errors stemmed from the models’ difficulties in comprehending study design as per the inclusion criteria, with some errors also arising from the lack of sufficient information in titles and abstracts needed for decision-making. All three models displayed a high proportion of contextual incoherence and misunderstanding the questions with minor incidences of clinical cognitive biases, such as confirmation bias and hallucination, which were particularly predominant with Gemini-P. Including prompts with detailed guidance on identifying phase III studies, along with the BoDapproach, could mitigate these issues. In such cases, a human-in-the-loop approach would allow final decisions on borderline studies, further enhancing accuracy and reducing relevant exclusions

The current study has several strengths. First, this study is the first to evaluate a collaborative framework that simulates a two-human-reviewers setting, closely aligning with real-world practices and enhancing the reliability and validity of the automated screening process. Second, we utilized five real-world clinical data-sets comprising over 10,000 articles, each annotated by two independent reviewers. To enhance fidelity, a third reviewer cross-checked decision labels across all articles in the five datasets. These fully annotated datasets, along with machine-generated decision labels and explanations, have been made publicly available, providing a valuable resource for further research and future development efforts in this space. Third, we conducted a formalized error analysis led by experienced systematic reviewers, categorizing mistakes using a predefined schema to gain deeper insights into specific areas where the models tend to make mistakes. Fourth, we used SOTA models with zero-shot prompts to make decisions without requiring extensive pre-training on specific systematic review datasets or in-context exemplars, making our proposed collaborative approach adaptable and easily applicable to systematic reviews synthesizing evidence for other clinical questions. Finally, we also explored the potential of LLM-LLM conversational interactions, presenting a preliminary case study on a novel mechanism for conflict resolution and decision refinement. This method highlights the capability of advanced LLMs to engage in meaningful dialogue, leading to more accurate and consensus-driven outcomes.

This study introduces a promising framework for leveraging collaborative LLMs to automate the screening phase of systematic reviews, effectively simulating the dual-reviewer model, conversational LLM interaction and demonstrating high precision, recall, and substantial workload reduction The findings suggest that LLM collaboration, with structured conflict resolution, could enhance the accuracy and consistency of evidence synthesis process, particularly for living guidelines. However, the variability in model performance highlights the need for deeper understanding of model reasoning. This work lays a strong foundation for future research into scalable, trustworthy, and adaptive LLMs-based systems in clinical evidence synthesis workflows, inviting further exploration of how best to integrate LLMs into real-world systematic review pipelines.

## Funding Source

This work was supported by the National Institute of Health U24 grant (U24CA265879-01-1) and Carolyn Ann Kennedy Bacon Fund.

## IRB Approval

IRB approval was deemed exempt given that this work relied on publically available publications (title and abstracts) and did not include any human or animal related data.

## Acknowledgements

None

## Notes

### Competing Interest Statement

The authors have declared no competing interest.

### Funding Statement

This study was funded by NIH (U24CA265879-01-1) and Carolyn-Ann-Kennedy-Bacon Fund

## REFERENCES

1. Chai KEK, Lines RLJ, Gucciardi DF, Ng L. Research Screener: a machine learning tool to semi-automate abstract screening for systematic reviews. Syst Rev. 2021;10(1):93.

2. Polanin JR, Pigott TD, Espelage DL, Grotpeter JK. Best practice guidelines for abstract screening large-evidence systematic reviews and meta-analyses. Res Synth Methods. 2019;10(3):330–342.

3. Stoll CRT, Izadi S, Fowler S, Green P, Suls J, Colditz GA. The value of a second reviewer for study selection in systematic reviews. Res Synth Methods. 2019;10(4):539–545.

4. van de Schoot R, de Bruin J, Schram R, et al. An open source machine learning framework for efficient and transparent systematic reviews. Nat Mach Intell. 2021;3(2):125–133.

5. Sandner E, Gütl C, Jakovljevic I, Wagner A. Screening automation in systematic reviews: analysis of tools and their machine learning capabilities. Stud Health Technol Inform. 2024;313:179–185.

6. Brown TB, Mann B, Ryder N, et al. Language models are few-shot learners. arXiv. 2020. Accessed July 20, 2025. https://arxiv.org/abs/2005.14165

7. Cao C, Sang J, Arora R, et al. Development of prompt templates for large language model–driven screening in systematic reviews. Ann Intern Med. 2025;178(3):389–401.

8. Khan MA, Ayub U, Naqvi SAA, et al. Collaborative large language models for automated data extraction in living systematic reviews. J Am Med Inform Assoc. 2025;32(4):638–647.

9. Riaz IB, Naqvi SAA, Hasan B, Murad MH. Future of evidence synthesis: automated, living, and interactive systematic reviews and meta-analyses. Mayo Clin Proc Digit Health. 2024;2(3):361–365.

10. Riaz IB, Naqvi SAA, He H, et al. First-line systemic treatment options for metastatic castration-sensitive prostate cancer: a living systematic review and network meta-analysis. JAMA Oncol. 2023;9(5):635–645.

11. Naqvi SAA, Anjum MU, Bibi A, et al. Systemic treatment options for metastatic castration-resistant prostate cancer: a living systematic review. medRxiv. Preprint posted online 2025. doi:10.1101/2025.01.01.23298765

12. Riaz IB, He H, Ryu AJ, et al. A living, interactive systematic review and network meta-analysis of first-line treatment of metastatic renal cell carcinoma. Eur Urol. 2021;80(6):712–723.

13. Sonbol MB, Riaz IB, Naqvi SAA, et al. Systemic therapy and sequencing options in advanced hepatocellular carcinoma: a systematic review and network meta-analysis. JAMA Oncol. 2020;6(12):e204930.

14. Kojima T, Gu SS, Reid M, et al. Large language models are zero-shot reasoners. arXiv. 2022. Accessed July 20, 2025. https://arxiv.org/abs/2205.11916

15. Achiam OJ, Adler S, Agarwal S, et al. GPT-4 technical report. OpenAI; 2023. Accessed July 20, 2025. https://arxiv.org/abs/2303.08774

16. Anthropic. The Claude 3 model family: Opus, Sonnet, Haiku. 2023. Accessed July 20, 2025. https://www.anthropic.com/index/claude-3

17. Team G, Anil R, Borgeaud S, et al. Gemini: a family of highly capable multimodal models. arXiv. 2023. Accessed July 20, 2025. https://arxiv.org/abs/2312.11805

18. Kusa W, Lipani A, Knoth P, Hanbury A. An analysis of work saved over sampling in the evaluation of automated citation screening in systematic literature reviews. Intell Syst Appl. 2023;18:200193.

19. Ouzzani M, Hammady H, Fedorowicz Z, Elmagarmid A. Rayyan—a web and mobile app for systematic reviews. Syst Rev. 2016;5(1):210.

20. Gates A, Johnson C, Hartling L. Technology-assisted title and abstract screening for systematic reviews: a retrospective evaluation of the Abstrackr machine learning tool. Syst Rev. 2018;7(1):45.

21. Syriani E, David I, Kumar G. Screening articles for systematic reviews with ChatGPT. J Comput Lang. 2024;80:101287.

22. Robinson A, Thorne W, Wu B, et al. Bio-SIEVE: exploring instruction tuning large language models for systematic review automation. arXiv. 2023. Accessed July 20, 2025. https://arxiv.org/abs/2308.06610

23. Wang S, Scells H, Zhuang S, et al. Zero-shot generative large language models for systematic review screening automation. In: Lecture Notes in Computer Science. Cham: Springer; 2024:403–420.

24. Martínez García L, Sanabria AJ, García Alvarez E, et al. The validity of recommendations from clinical guidelines: a survival analysis. CMAJ. 2014;186(16):1211–1219.

25. Densen P. Challenges and opportunities facing medical education. Trans Am Clin Climatol Assoc. 2011;122:48–58.

26. Strobl MAR, Gallaher J, Robertson-Tessi M, et al. Treatment of evolving cancers will require dynamic decision support. Ann Oncol. 2023;34(10):867–884.

27. Pang T, Amul GGH. Rapid guidelines: timely and important guidance needed for setting standards and best practices. Health Res Policy Syst. 2018;16(1):56.

