## Supplemental Material for "Collaborative large language models (LLMs) are all you need for screening in systematic reviews"

**Online only supplementary material**

**Supplement Methods.** Detailed methods

**Supplement Figure 1**. Inference flow for each of the three models

**Supplement Figure 2**. Illustration of cross-LLM interaction for conflict resolution

**Supplement Figure 3**. Schema for error analysis

**Supplement Table 1.** Initial prompt used for main experiments

**Supplement Table 2.** Prompt used for conflict resolution

**Supplement Table 3.** All prompts used for LLM-LLM Interactions

**Supplement Table 4.** Example of LLM-LLM interaction for conflict resolution

**Supplement Table 5.** Distribution of errors for each independent individual large language model

**Supplement Table 6.** Error analysis for responses by GPT-4T

**Supplement Table 7.** Error analysis for responses by Claude-3S

**Supplement Table 8.** Error analysis for responses by Gemini-P

**Supplement Table 9.** Summary of error analyses across different projects by different models

**Supplementary Methods.** Detailed methods

**Literature search**

For each systematic review project, a systematic search was initially manually executed in Ovid MEDLINE and EMBASE to retrieve a list of citations relevant to each systematic review project. Subsequently, a living ‘auto’ search was implemented to retrieve new citations at pre-specified regular intervals using the Python packages “scrapy” and “requests”. The list of all citations was pre-processed to remove duplicates. Citations were assigned a unique identification number and converted into a predefined data schema for storage in a MySQL-based central data repository

**Manual annotation**

Citations are presented in a web-assisted graphical user interface where two independent reviewers screened citations based on prespecified inclusion/exclusion criteria for potential eligibility. The title and abstract of each citation are screened, and each citation is labeled based on the decision for potential inclusion or exclusion. For example, a citation deemed ineligible by title and abstract examination was labeled as “excluded by title/abstract.” Third, the full text of the citations, deemed eligible for inclusion by title and abstract screening, were reviewed for final inclusion in the systematic review. Each citation at the full-text review stage was then labeled in accordance with the decision made for inclusion or exclusion.

**Data processing**

The user-generated labels, along with the corresponding meta-data, and free text for each stage of study selection, were structured and organized in a central data repository and subsequently parsed into a JavaScript object notation (JSON) file. Decision labels for articles by each stage were transformed into a single binary variable with two levels: included (included by full-text review) and excluded (excluded by title/abstract or excluded by full-text review).

**Experimental setting**

The process of screening was formulated as a generative approach. Let us consider a set of study instances S corresponding to a systematic review dataset. In this set, the i^th instance is represented as S_i = {(t_i,a_i)} where t_i represents the title and a_i represents the abstract of the i^th study. Each title and abstract pair are evaluated based on predefined PICOS criteria (excluding the control criterion), and the study is included if four out of the remaining criteria (Population, Intervention, Outcome, Study Design) are met.

We evaluated three commercial (closed-source) LLMs on Zero-shot-CoT and proposed methods: (1) GPT-4-Turbo, (2) Google Gemini-Pro, and (3) Claude-3-Sonnet. We evaluate each model by accessing their APIs from respective providers. The evaluation is conducted on the versions of these models released in May 2024. It’s important to note that these models are regularly updated, so when reproducing the results, there is a possibility of variations.

Each model is evaluated at temperature 0 to ensure deterministic behavior and the maximum output of 1000 tokens. For the LLM-LLM interaction method, we use a temperature of 0.5 to ensure more liberty in generation capabilities during conversing with each other.

**Conversational interaction between two models**

In this approach, we developed a multi-agent system leveraging interactions between two advanced and best-performing LLMs, GPT-4 and Claude-3, to automate the systematic review process. First, each model is provided with a standardized system message instructing them to assess the studies' titles and abstracts and produce a reasoning chain for their inclusion or exclusion decisions. Initially, both models independently evaluated the study and generated their reasoning chains. If there was a conflict between their independent decisions, then these chains were combined into a comprehensive message, which was iteratively fed back to both models to refine their decisions through a structured conversation. This iterative process aimed to consolidate the insights from both models. The results, including the models' reasoning and final decisions, were systematically recorded and analyzed to assess the system's performance. This approach demonstrates the potential of using collaborative interactions between multiple LLMs to improve the efficiency and effectiveness of systematic reviews.

**Error analysis**

A systematic error analysis was performed by a trained systematic reviewer to assess the reasoning chain behind each model’s decision for each article.

The process began with a review of the gold labels, where the titles and abstracts of each study were re-examined to ensure consistency with the pre-defined PICOS criteria. Explanations provided by the models were then analyzed to understand deviations from human judgment. Errors were first stratified by the population, intervention, control/comparator, outcomes and study design (PICOS) elements, and further categorized using a pre-specified error schema to differentiate errors likely due to comprehension from those attributable to reasoning, as illustrated in **Figure 1.**

Comprehension Errors

Comprehension errors involved failures in understanding the context or content of the study, often manifesting as contextual incoherence. These were further analyzed for specific cognitive biases which include:

1) Misunderstanding the question: Did the model extract irrelevant information because it did not understand the question?
2) Redundancy: Did the model extract multiple redundant answers?
3) Hallucination/Confabulation: Did the model extract information not present in the input?

Reasoning Errors

Reasoning errors involved flaws in the logical process. These were subdivided into false association (incorrectly linking two pieces of information) or missing step (lacking key information to reach an answer). Reasoning errors were also further analyzed for the presence of cognitive biases:

1) Factual Error: Did the model make a factually incorrect assertion?
2) Anchoring Bias: Did the model focus on eligibility information early in the text, failing to incorporate further contradictory information?
3) Confirmation Bias: Did the model only acknowledge evidence in support of a decision, failing to look for contradictory information?
4) Premature Closure: Did the model reach a conclusion with insufficient evidence?
5) Omission Error: Did the model ignore or not acknowledge a pre specific component of PICOS?

Following the error categorization, detailed written explanations were provided to clarify the specific mistakes made by each model.

**Supplement Figure 1**. Inference flow for each of the three models

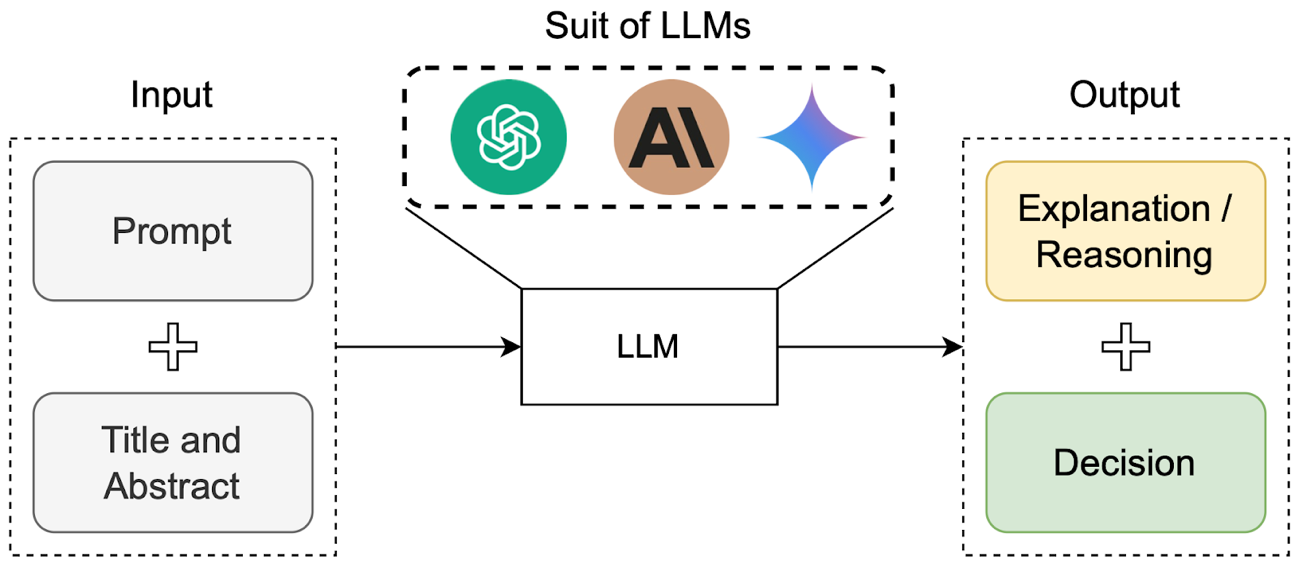

This figure illustrates how each model in this study was evaluated independently in a zero-shot setting.

**Supplementary Figure 2**. Illustration of cross-LLM interaction for conflict resolution

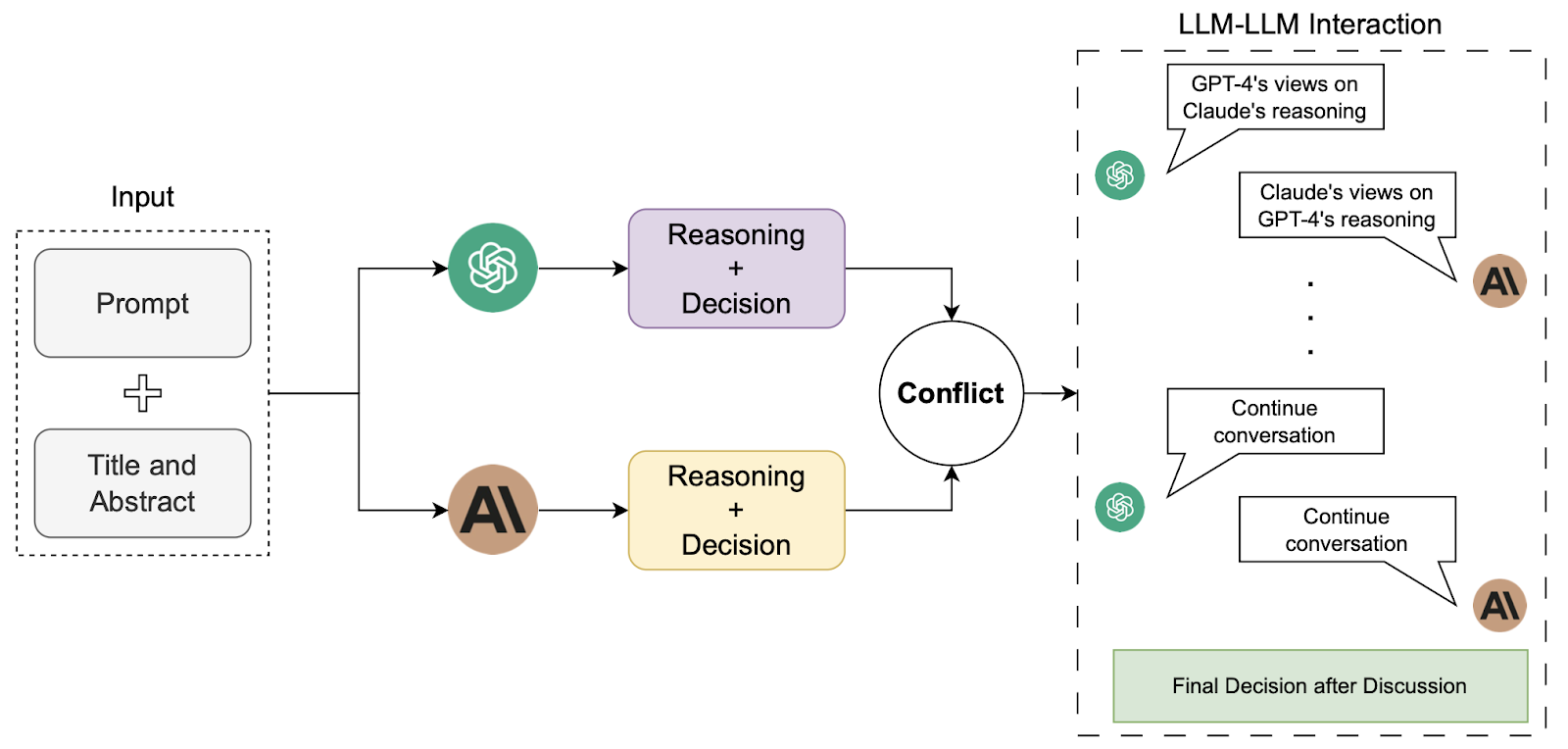

This figure illustrates that when a conflict arises between two models, they engage in an LLM–LLM interaction and continue discussing until they reach a final decision.

**Supplementary Figure 3**. Schema for error analysis

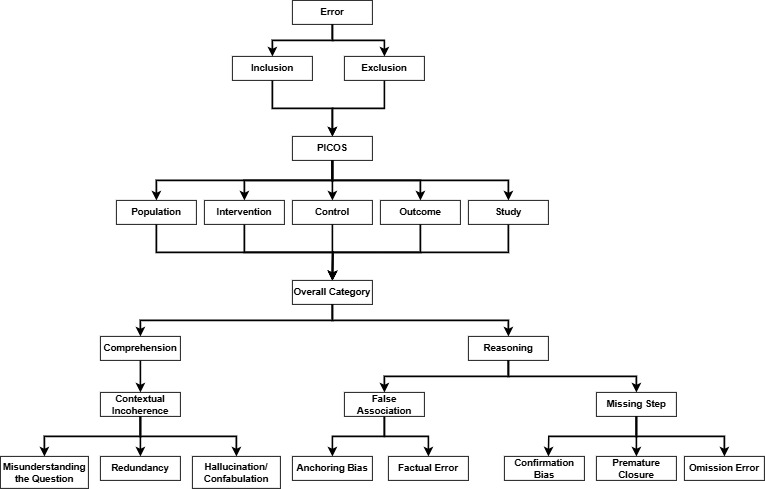

**Supplementary Table 1.** Initial prompt used for main experiments

**Initial Prompt:**

You are a systematic review expert. You are required to screen a given study by reading the title and abstract of the given study. You will decide based on the [component 1, component 2, component N] of the PICOS (population, intervention, control, outcome, study design] criteria. You will only include the given study if all of the criteria are met/satisfied. Please produce only one label for the given study: "include", or "exclude". Please also provide a step-by-step explanation for your decision. DO NOT generate anything apart from Include or Exclude for Decision. Always provide a Decision after an explanation.

FORMAT:

Explanation: [step-by-step explanation]

Decision: Include/Exclude

PICOS:

<Dataset specific PICOS criteria>

Title: <add title text>

Abstract: <add abstract text>

For the main experiments in Figure 1 (main paper), we use the same prompt given above for all methods. Specifically, each method begins with this identical instruction as its prompt.

**Supplementary Table 2.** Prompt used for conflict resolution

**Prompt:**

You are a systematic review expert. You are required to screen a given study by reading the title and abstract of the given study. You will decide based on the [component 1, component 2, component N] of the PICOS (population, intervention, control, outcome, study design] criteria. Since there is a conflict between two models in making decision, you are also given their reasoning chains and label for you to analyze. Based on all this, you will only include the given study if all of the criteria are met/satisfied. Please produce only one label for the given study: "include", or "exclude". Please also provide a step-by-step explanation for your decision. DO NOT generate anything apart from Include or Exclude for Decision. Always provide a Decision after an explanation.

FORMAT:

Explanation: [step-by-step explanation]

Decision: Include/Exclude

PICOS:

<Dataset specific PICOS criteria>

Title: <add title text>

Abstract: <add abstract text>

Reasoning Chain for GPT-4: <add reasoning chain text>

Reasoning Chain for Claude-3: <add reasoning chain text>

**Supplementary Table 3.** All prompts used for LLM-LLM Interactions

**System Prompt:**

You are a systematic review expert. You are required to screen a given study by reading the title and abstract of the given study. You will decide based on the [component 1, component 2, component N] of the PICOS (population, intervention, control, outcome, study design] criteria. Out of them, please ignore the control criteria. You will only include the given study if four of the criteria (population, intervention, outcome, study design) are met/satisfied. Please produce only one label for the given study: "include", or "exclude". Please provide a reasoning chain for your answer.

**Initial Prompt:**

Hey, we need to review this article to make final decision based on PICOS. After producing an independent decision label for a given study, we will compare our decision labels and reach a final decision. If our decisions are the same, we will finalize that decision. If our decisions are different, then we will review each other's reasoning and reach a consensus for the final decision.

PICOS:

<Dataset specific PICOS criteria>

FORMAT:

Explanation: [step-by-step explanation]

Decision: Include/Exclude

**Prompt for Starting Conversation:**

Based on the provided title and abstract, you will notice that there are disagreements in the predictions made by you and Claude-3. Please engage in a discussion with Claude-3 to identify and resolve any disagreements regarding the PICO criteria. Claude-3, please present your arguments as well. Continue the conversation through multiple rounds until you reach a consensus. Once a final decision is made, ALWAYS present it in the following format: 'Final Decision: Include/Exclude' with the selected label. Additionally, I have included the reasoning chains from both you (GPT-4) and Claude-3, along with the title and abstract to begin with.

Title: <add title text>

Abstract: <add abstract text>

Reasoning Chain for GPT-4: <add reasoning chain text>

Reasoning Chain for Claude-3: <add reasoning chain text>

Each model participating in conversation receives a **“System Prompt”** instructing it to act as a systematic reviewer. They then receive an **“Initial Prompt”** to produce their first independent decision. If the models’ decisions conflict, the **“Prompt for Starting Conversation”** is invoked to initiate a discussion among them.

**Supplementary Table 4.** Example of LLM-LLM interaction for conflict resolution

**User:**

Based on the provided title and abstract, you will notice that there are disagreements in the predictions made by you and Claude-3. Please engage in a discussion with Claude-3 to identify and resolve any disagreements regarding the PICO criteria. Claude-3, please present your arguments as well. Continue the conversation through multiple rounds until you reach a consensus. Once a final decision is made, ALWAYS present it in the following format: 'Final Decision: Include/Exclude' with the selected label. Additionally, I have included the reasoning chains from both you (GPT-4) and Claude-3, along with the title and abstract to begin with.

**Input:**

Title: Single-agent therapy with bicalutamide: a comparison with medical or surgical castration in the treatment of advanced prostate carcinoma

Abstract: OBJECTIVES: Single-agent therapy with bicalutamide, a nonsteroidal antiandrogen, was compared with castration, either surgical METHODS: In an open, randomized, multicenter trial, patients were randomized to treatment with 50 mg bicalutamide (n = 243) once daily or to castration (n = 243), either orchiectomy or depot injection of goserelin acetate every 28 days. Primary efficacy endpoints were times to treatment failure and objective disease progression and survival. Assessments included review of measurable metastases, prostate dRESULTS: The median duration of therapy was 39 weeks for bicalutamide-treated patients and 42 weeks for castrated patients; treatment failure occurred in 53% and 42% and disease progression in 43% and 33%, respectively. Treatment effects favored castration for both endpoints (P < or = 0.002), with hazard ratios (bicalutamide:castration) of 1.54 (95% confidence interval [CI], 1.18 to 2.00) for time to treatment failure and 1.6 (95% CI, 1.19 to 2.15) for time to disease progression. From the 1-year survival analysis, the hazard ratio for probability of death was 1.29 (95% CI, 0.96 to 1.72). Thus far, with a median follow-up of 86 weeks, median survival has not been reached in either group. Changes from baseline in several quality of life variables were significantly different (P < or = 0.01) between treatment groups periodically from months 1 to 6, and all favored bicalutamide. Overall, the antiandrogen was well tolerated compared with castCONCLUSIONS: Although a dosage of 50 mg of bicalutamide once daily was not as effective as castration, the favorable quality of life outcomes and the low incidence of nonhormonal adverse events provide reasons to evaluate bicalutamide, as a single therapeutic agent, at higher doses.

**GPT-4 Reasoning Chain:**

Explanation: The population in the study is patients with advanced prostate cancer, which aligns with the PICOS criteria of metastatic castration/hormone resistant/refractory prostate cancer. The intervention in the study is single-agent therapy with bicalutamide, a systemic treatment option without surgery/radiation, which aligns with the PICOS criteria. The control in the study is castration, either surgical or medical, which can be considered as a standard of care for the population. The study design is a randomized, multicenter trial, which is a type of Phase III randomized controlled clinical trial. Therefore, all the PICOS criteria are met in this study.

Decision: Include

**Claude-3 Reasoning Chain:**

Explanation:

1. The study population is patients with untreated Stage D2 prostate cancer, which meets the criterion of metastatic prostate cancer.

2. The intervention is single-agent therapy with bicalutamide, a systemic treatment option without surgery/radiation, which satisfies the intervention criterion.

3. The control group underwent castration, either surgical (orchiectomy) or medical (goserelin acetate), which can be considered the standard of care for this population.

4. The study design is an open, randomized, multicenter trial, which is a randomized controlled clinical trial. However, it is not explicitly stated as a Phase III trial.

Decision: Exclude

The study meets most of the PICOS criteria, including the population, intervention, and control. However, the study design is not explicitly stated as a Phase III trial, which is a requirement according to the provided criteria. Therefore, the study should be excluded from the systematic review.

**Conversation:**

**GPT-4 (First Response):**

In reviewing the reasoning chains from both GPT-4 and Claude-3, it's clear there is a disagreement specifically concerning the study design criterion. Let's analyze and discuss this point to reach a consensus.

**GPT-4 Argument:**

The study design is identified as a randomized, multicenter trial. GPT-4 assumes this to be a Phase III randomized controlled clinical trial based on the methodology described, which generally aligns with the characteristics of Phase III trials in terms of comparing a new treatment to the standard of care in a large group of participants.

**Claude-3 Argument:**

Claude-3 specifically points out that the study design is not explicitly stated as a Phase III trial. This omission is crucial for Claude-3's decision to exclude the study, as the explicit identification of the trial phase is a requirement according to the criteria set forth.

**Discussion:**

The primary point of contention lies in whether the explicit designation of the trial as "Phase III" is necessary for inclusion. While it is true that the study is a randomized, multicenter trial comparing a new treatment against a standard treatment, the lack of explicit mention as a Phase III trial could be problematic if strict adherence to terminology is required for systematic review inclusion criteria.

However, considering the broader context of clinical research, Phase III trials are typically designed to confirm the efficacy of a treatment compared to the standard of care in a larger patient group, which aligns with the description of the study design provided in the abstract. Therefore, the assumption that this study could be a Phase III trial based on its characteristics is reasonable.

**Conclusion:**

Given that the study design described corresponds closely with what is typically expected of a Phase III trial, and considering the importance of the other met criteria (population, intervention, outcome), it seems appropriate to lean towards inclusion, provided that the systematic review does not strictly require the explicit mention of "Phase III" as part of the study design description.

Final Decision: Include

**Claude-3 (Response to GPT-4):**

Thank you for providing the reasoning chains from both GPT-4 and Claude-3, as well as the title and abstract of the study. Let's engage in a discussion to resolve the disagreement regarding the study design criterion and reach a consensus.

**GPT-4's Argument:**

You make a valid point that the study design, described as a randomized, multicenter trial, aligns with the characteristics of a Phase III randomized controlled clinical trial. The methodology of comparing a new treatment (bicalutamide) to a standard of care (castration) in a large group of participants is indeed typical of Phase III trials.

**Claude-3's Argument:**

Claude-3 raises an important concern that the study design is not explicitly stated as a Phase III trial in the abstract. Strict adherence to the inclusion criteria would require the trial phase to be clearly specified.

**Discussion:**

While I understand Claude-3's perspective on the importance of explicitly stating the trial phase, I believe that the study design described in the abstract strongly suggests that this is a Phase III trial. The key characteristics, such as the randomized, multicenter nature of the trial, the comparison of a new treatment to a standard of care, and the large sample size (486 patients in total), are all indicative of a Phase III trial.

Moreover, the purpose of the systematic review should be considered. If the review aims to comprehensively evaluate the efficacy of bicalutamide in the treatment of advanced prostate cancer, excluding this study based on the technicality of not explicitly stating "Phase III" could potentially omit valuable data.

However, if the systematic review protocol strictly requires the explicit mention of "Phase III" in the study design description, then Claude-3's argument for exclusion would be valid.

**Conclusion:**

After considering both arguments and the context of the systematic review, I propose that we include this study in the review. The study design described in the abstract is highly suggestive of a Phase III trial, and the study meets all other relevant PICO criteria (population, intervention, outcome). Including this study would contribute to a more comprehensive evaluation of bicalutamide's efficacy in treating advanced prostate cancer.

Claude-3, I hope this discussion helps clarify my perspective. Please let me know if you have any further thoughts or concerns regarding this decision.

Final Decision: Include

**GPT-4 (Follow-up):**
Final Decision: Include

The study meets the necessary criteria for inclusion based on its design, population, intervention, and outcomes. Although the study design is not explicitly labeled as a Phase III trial, the characteristics described align well with those typically associated with Phase III trials. This includes a large, randomized, multicenter format that compares a new treatment against a standard of care. Given the importance of the information this study provides in assessing the efficacy and quality of life outcomes of bicalutamide treatment in advanced prostate cancer, it is appropriate to include it in the systematic review.

**Claude-3 (Final Response):**

Thank you for the thoughtful discussion, Claude-3. After considering both of our arguments and the context of the systematic review, I agree that we should include this study in the review.

Your point about the study design not being explicitly labeled as a Phase III trial is valid. However, as I mentioned, the characteristics of the study, such as its randomized, multicenter nature, large sample size, and comparison of a new treatment to a standard of care, strongly suggest that it is a Phase III trial.

Moreover, the purpose of the systematic review is to comprehensively evaluate the efficacy of bicalutamide in the treatment of advanced prostate cancer. Excluding this study based on the technicality of not explicitly stating "Phase III" could potentially omit valuable data that contributes to this goal.

Given that the study meets all other relevant PICO criteria (population, intervention, outcome) and its design is highly suggestive of a Phase III trial, I believe including it in the review is the most appropriate decision.

Thank you for engaging in this discussion and helping to ensure that we make a well-reasoned decision.

Final Decision: Include

This shows a sample dialogue between GPT-4 (green) and Claude 3 (yellow) during conflict resolution. After exchanging reasoning, Claude 3 revises its initial stand and ultimately agrees with GPT-4 on including the article.

**Supplementary Table 5.** Distribution of errors for each independent individual large language model

| **Number of errors** | **mHSPC** | | **mCRPC** | | **RCC** | | **mHCC** | | **PARPi Toxicity** | |
| --- | --- | --- | --- | --- | --- | --- | --- | --- | --- | --- |
|  | **N = 69** | | **N = 257** | | **N = 385** | | **N = 272** | | **N = 1294** | |
| ***Error at the level of each independent individual large language models - N (%)*** | | | | | | | | | | |
| GPT-4T | 15 (21.73) | | 58 (22.57) | | 147 (38.18) | | 136 (50) | | 161 (12.44) | |
| Claude-3S | 13 (18.84) | | 59 (22.95) | | 103 (26.75) | | NA* | | 315 (24.34) | |
| Gemini-P | 41 (59.42) | | 140 (54.47) | | 135 (35.06) | | 136 (50) | | 818 (63.21) | |
| ***Errors by false decision at the level of each independent individual large language models - N (%)*** | | | | | | | | | | |
|  | **Exclusion** | **Inclusion** | **Exclusion** | **Inclusion** | **Exclusion** | **Inclusion** | **Exclusion** | **Inclusion** | **Exclusion** | **Inclusion** |
| GPT-4T | 1 (6.7) | 14 (93.3) | 5 (8.6) | 53 (91.4) | 2 (1.4) | 145 (98.6) | 0 (0) | 136 (100) | 33 (20) | 128 (80) |
| Claude-3S | 0 (0) | 13 (100) | 13 (22) | 46 (78) | 1 (1.0) | 102 (99) | NA* | NA* | 7 (2.2) | 308 (97.8) |
| Gemini-P | 3 (7.3) | 38 (92.7) | 18 (13) | 122 (87) | 23 (17) | 112 (83) | 1 (0.7) | 135 (99.3) | 57 (7.0) | 761 (93) |

Abbreviations: mHSPC: metastatic hormone-sensitive prostate cancer; mCRPC: metastatic castration resistant prostate cancer; RCC: renal cell carcinoma; mHCC: metastatic hepatocellular carcinoma; PARPi: poly (ADP-ribose) polymerase inhibitors; GPT-4T: generative pre-trained transformer; Claude-3S: Claude-3-Sonnet; GeminiP: Gemini-Pro

*Claude-3S was not evaluated on the mHCC dataset.

**Supplementary Table 6.** Error analysis for responses by GPT-4T

| **GPT-4T** | **mHSPC** | **mCRPC** | **RCC** | **mHCC** | **PARPi Toxicity** | **Total** |
| --- | --- | --- | --- | --- | --- | --- |
| Exclude | 1 / 15 (6.7%) | 5 / 58 (8.6%) | 2 / 147 (1.4%) | 0 / 136 (0%) | 33 / 161 (20%) | 41/517 (7.93%) |
| Include | 14 / 15 (93.3%) | 53 / 58 (91.4%) | 145 / 147 (98.6%) | 136 / 136 (100%) | 128 / 161 (80%) | 476/517 (92.07%) |
| ***PICOS Category*** |  |  |  |  |  |  |
| Population | 1 / 15 (6.7%) | 20 / 58 (34%) | 34 / 147 (23.1%) | 4 / 136 (3.0%) | 26 / 161 (16.1%) | 85/517 (16.44%) |
| Intervention | 3 / 15 (20%) | 9 / 58 (16%) | 13 / 147 (8.8%) | 19 / 136 (14%) | 7 / 161 (4.3%) | 51/517 (9.86%) |
| Control | 0 / 15 (0%) | 0 / 58 (0%) | 2 / 147 (1.4%) | 0 / 136 (0%) | 2 / 161 (1.2%) | 4/517 (0.77%) |
| Outcome | 0 / 15 (0%) | 2 / 58 (3.0%) | 0 / 147 (0%) | 0 / 136 (0%) | 11 / 161 (6.8%) | 13/517 (2.51%) |
| Study | 11 / 15 (73.3%) | 27 / 58 (47%) | 98 / 147 (66.7%) | 113 / 136 (83%) | 115 / 161 (71.4%) | 364/517 (70.40%) |
| ***Overall Category: Comprehension or Reasoning*** |  |  |  |  |  |  |
| Comprehension | 14 / 15 (93.3%) | 47 / 58 (81%) | 143 / 147 (97.3%) | 136 / 136 (100%) | 156 / 161 (96.9%) | 496/517 (95.93%) |
| Insufficient Information provided in the abstract | 1 / 15 (6.7%) | 11 / 58 (19%) | 4 / 147 (2.7%) | 0 / 136 (0%) | 5 / 161 (3.1%) | 21/517 (4.06%) |
| ***LLM Reasoning Mistake*** |  |  |  |  |  |  |
| Contextual Incoherence | 14 / 14 (100%) | 47 / 47 (100%) | 143 / 143 (100%) | 136 / 136 (100%) | 156 / 156 (100%) | 496/496 (100%) |
| ***Clinical Cognitive Bias Correlate*** |  |  |  |  |  |  |
| Misunderstanding the question | 14 / 14 (100%) | 47 / 47 (100%) | 143 / 143 (100%) | 136 / 136 (100%) | 156 / 156 (100%) | 496/496 (100%) |

Abbreviations: mHSPC: metastatic hormone-sensitive prostate cancer; mCRPC: metastatic castration resistant prostate cancer; RCC: renal cell carcinoma; mHCC: metastatic hepatocellular carcinoma; PARPi: poly (ADP-ribose) polymerase inhibitors; GPT-4T: generative pre-trained transformer; Claude-3S: Claude-3-Sonnet; GeminiP: Gemini-Pro

**Supplementary Table 7.** Error analysis for responses by Claude-3S

| **Claude-3S** | **mHSPC** | **mCRPC** | **RCC** | **PARPi Toxicity** | **Total** |
| --- | --- | --- | --- | --- | --- |
| Exclude | 0 / 13 (0%) | 13 / 59 (22%) | 1 / 103 (1.0%) | 7 / 315 (2.2%) | 21/490 (4.29%) |
| Include | 13 / 13 (100%) | 46 / 59 (78%) | 102 / 103 (99%) | 308 / 315 (97.8%) | 469/490 (95.71%) |
| ***PICOS Category*** |  |  |  |  |  |
| Population | 0 / 13 (0%) | 11 / 59 (19%) | 17 / 102 (16.7%) | 0 / 298 (0%) | 28/472 (5.93%) |
| Intervention | 2 / 13 (15.4%) | 12 / 59 (20%) | 6 / 102 (5.9%) | 211 / 298 (70.8%) | 231/472 (48.94%) |
| Control | 1 / 13 (7.7%) | 6 / 59 (10%) | 2 / 102 (1.9%) | 0 / 298 (0%) | 9/472 (1.90%) |
| Outcome | 0 / 13 (0%) | 0 / 59 (0%) | 0 / 102 (0%) | 16 / 298 (5.4%) | 16/472 (3.39%) |
| Study | 10 / 13 (76.9%) | 30 / 59 (51%) | 77 / 102 (75.5%) | 71 / 298 (23.8%) | 188/472 (39.83%) |
| ***Overall Category: Comprehension or Reasoning*** |  |  |  |  |  |
| Comprehension | 11 / 13 (85%) | 43 / 59 (73%) | 99 / 103 (96.1%) | 296 / 315 (94%) | 449/490 (91.63%) |
| Insufficient information provided in the abstract | 2 / 13 (15%) | 16 / 59 (27%) | 4 / 103 (3.9%) | 19 / 315 (6.0%) | 41/490 (8.36%) |
| ***LLM Reasoning Mistake*** |  |  |  |  |  |
| Contextual Incoherence | 11 / 11 (100%) | 43 / 43 (100%) | 99 / 99 (100%) | 296 / 296 (100%) | 449/449 (100%) |
| ***Clinical Cognitive Bias Correlate*** |  |  |  |  |  |
| Misunderstanding the question | 11 / 11 (100%) | 43 / 43 (100%) | 99 / 99 (100%) | 296 / 296 (100%) | 449/449 (100%) |

Abbreviations: mHSPC: metastatic hormone-sensitive prostate cancer; mCRPC: metastatic castration resistant prostate cancer; RCC: renal cell carcinoma; PARPi: poly (ADP-ribose) polymerase inhibitors; GPT-4T: generative pre-trained transformer; Claude-3S: Claude-3-Sonnet; GeminiP: Gemini-Pro

**Supplementary Table 8.** Error analysis for responses by Gemini-P

| **Gemini-P** | **mHSPC** | **mCRPC** | **RCC** | **mHCC** | **PARPi Toxicity** | **Total** |
| --- | --- | --- | --- | --- | --- | --- |
| Exclude | 3 / 41 (7.3%) | 18 / 140 (13%) | 23 / 135 (17%) | 1 / 136 (0.7%) | 57 / 818 (7.0%) | 102/1270 (8.03%) |
| Include | 38 / 41 (92.7%) | 122 / 140 (87%) | 112 / 135 (83%) | 135 / 136 (99.3%) | 761 / 818 (93%) | 1168/1270 (91.96%) |
| ***PICOS Category*** |  |  |  |  |  |  |
| Population | 13 / 41 (31.7%) | 28 / 140 (20%) | 38 / 134 (28.4%) | 21 / 136 (15.4%) | 22 / 796 (2.8%) | 122/1247 (9.78%) |
| Intervention | 9 / 41 (22%) | 27 / 140 (19.3%) | 9 / 134 (6.7%) | 23 / 136 (16.9%) | 649 / 796 (81.5%) | 717/1247 (57.49%) |
| Control | 2 / 41 (4.9%) | 11 / 140 (7.9%) | 5 / 134 (3.7%) | 0 / 136 (0%) | 9 / 796 (1.1%) | 27/1247 (2.16%) |
| Outcome | 3 / 41 (7.3%) | 0 / 140 (0%) | 4 / 134 (2.9%) | 1 / 136 (0.7%) | 30 / 796 (3.8%) | 38/1247 (3.05%) |
| Study | 14 / 41 (34.1%) | 74 / 140 (52.8%) | 78 / 134 (58.2%) | 91 / 136 (66.9%) | 86 / 796 (10.8%) | 343/1247 (27.50%) |
| ***Overall Category: Comprehension or Reasoning*** |  |  |  |  |  |  |
| Comprehension | 39 / 41 (95.1%) | 118 / 140 (84%) | 132 / 135 (97.8%) | 136 / 136 (100%) | 791 / 818 (96.7%) | 1216/1270 (95.74%) |
| Reasoning | 0 / 41 (0%) | 0 / 140 (0%) | 0 / 135 (0%) | 0 / 136 (0%) | 4 / 818 (0.5%) | 4/1270 (0.31%) |
| Insufficient Information provided in the abstract | 2 / 41 (4.9%) | 22 / 140 (16%) | 3 / 135 (2.2%) | 0 / 136 (0%) | 23 / 818 (2.8%) | 50/1270 (3.93%) |
| ***LLM Reasoning Mistake*** |  |  |  |  |  |  |
| Contextual Incoherence | 39 / 39 (100%) | 118 / 118 (100%) | 132 / 132 (100%) | 136 / 136 (100%) | 791 / 795 (99.5%) | 1216/1220 (99.67%) |
| Missing Step | 0 / 39 (0%) | 0 / 118 (0%) | 0 / 132 (0%) | 0 / 136 (0%) | 4 / 795 (0.5%) | 4/1220 (0.32%) |
| ***Clinical Cognitive Bias Correlate*** |  |  |  |  |  |  |
| Confirmation Bias | 0 / 39 (0%) | 0 / 118 (0%) | 0 / 132 (0%) | 0 / 136 (0%) | 3 / 795 (0.4%) | 3/1220 (0.25%) |
| Hallucination/Confabulation | 0 / 39 (0%) | 0 / 118 (0%) | 0 / 132 (0%) | 7 / 136 (5.1%) | 0 / 795 (0%) | 7/1220 (0.57%) |
| Misunderstanding the question | 39 / 39 (100%) | 118 / 118 (100%) | 132 / 132 (100%) | 129 / 136 (94.9%) | 791 / 795 (99.5%) | 1209/1220 (99.09%) |
| Premature Closure | 0 / 39 (0%) | 0 / 118 (0%) | 0 / 132 (0%) | 0 / 136 (0%) | 1 / 795 (0.1%) | 1/1220 (0.08%) |

Abbreviations: mHSPC: metastatic hormone-sensitive prostate cancer; mCRPC: metastatic castration resistant prostate cancer; RCC: renal cell carcinoma; mHCC: metastatic hepatocellular carcinoma; PARPi: poly (ADP-ribose) polymerase inhibitors; GPT-4T: generative pre-trained transformer; Claude-3S: Claude-3-Sonnet; GeminiP: Gemini-Pro; Claude-3S: Claude-3-Sonnet; GeminiP: Gemini-Pro

**Supplementary Table 9.** Summary of error analyses across different projects by different models

| **Models** | ***Total Errors*** | | | | | | | | | |
| --- | --- | --- | --- | --- | --- | --- | --- | --- | --- | --- |
|  | **mHSPC** | | **mCRPC** | | **RCC** | | **mHCC** | | **PARP Toxicity** | |
|  | **69** | | **257** | | **385** | | **272** | | **1294** | |
| ***Errors by Each Model - N (%)*** | | | | | | | | | | |
| GPT-4T | 15 (21.73) | | 58 (22.57) | | 147 (38.18) | | 136 (50) | | 161 (12.44) | |
| Claude-3S | 13 (18.84) | | 59 (22.95) | | 103 (26.75) | | NA | | 315 (24.34) | |
| Gemini-P | 41 (59.42) | | 140 (54.47) | | 135 (35.06) | | 136 (50) | | 818 (63.21) | |
| ***Incorrect Exclusion or Inclusion by Each Model - n/N (%)*** | | | | | | | | | | |
|  | **Exclusion** | **Inclusion** | **Exclusion** | **Inclusion** | **Exclusion** | **Inclusion** | **Exclusion** | **Inclusion** | **Exclusion** | **Inclusion** |
| GPT-4T | 1 / 15 (6.7) | 14 / 15 (93.3) | 5 / 58 (8.6) | 53 / 58 (91.4) | 2 / 147 (1.4) | 145 / 147 (98.6) | 0 / 136 (0) | 136 / 136 (100) | 33 / 161 (20) | 128 / 161 (80) |
| Claude-3S | 0 / 13 (0) | 13 / 13 (100) | 13 / 59 (22) | 46 / 59 (78) | 1 / 103 (1.0) | 102 / 103 (99) | NA | NA | 7 / 315 (2.2) | 308 / 315 (97.8) |
| Gemini-P | 3 / 41 (7.3) | 38 / 41 (92.7) | 18 / 140 (13) | 122 / 140 (87) | 23 / 135 (17) | 112 / 135 (83) | 1 / 136 (0.7) | 135 / 136 (99.3) | 57 / 818 (7.0) | 761 / 818 (93) |

Abbreviations: mHSPC: metastatic hormone-sensitive prostate cancer; mCRPC: metastatic castration resistant prostate cancer; RCC: renal cell carcinoma; mHCC: metastatic hepatocellular carcinoma; PARPi: poly (ADP-ribose) polymerase inhibitors; GPT-4T: generative pre-trained transformer; Claude-3S: Claude-3-Sonnet; GeminiP: Gemini-Pro; Claude-3S: Claude-3-Sonnet; GeminiP: Gemini-Pro
